# Inadequate intention to receive Covid-19 vaccination: indicators for public health messaging needed to improve uptake in UK

**DOI:** 10.1101/2020.12.07.20243881

**Authors:** Patrik Bachtiger, Alexander Adamson, William A Maclean, Jennifer K Quint, Nicholas S Peters

## Abstract

Data promising effective Covid-19 vaccines have accelerated the UK’s mass vaccination programme. The UK public’s attitudes to the government’s prioritisation list are unknown, and achieving critical population immunity will require the remaining majority to accept both vaccination and the delay in access of up to a year or more. This cross-sectional observational study sent an online questionnaire to registrants of the UK National Health Service’s largest personal health record. Question items covered willingness for Covid-19 vaccine uptake and attitudes to prioritisation. Among 9,122 responses, 71.5% indicated wanting a vaccine, below what previous modelling indicated as critical levels for progressing towards herd immunity. 22.7% disagreed with the prioritisation list, though 70.3% were against being able to expedite vaccination through payment. Age and female gender were, respectively, strongly positively and negatively associated with wanting a vaccine. Teachers and Black, Asian and Minority Ethnic (BAME) groups were most cited by respondents for prioritisation. This study identifies factors to inform the public health messaging critical to improving uptake.

## Introduction

Data promising effective Covid-19 vaccines has accelerated the UK’s mass vaccination programme. The UK government’s prioritisation list is based only on age, care-home residency, preexisting disease, and health and care worker status.^1^ But achieving the critical level of population immunity will require the remaining majority to accept both vaccination and the delay of up to a year or more^2^ in them receiving it if an adequate level of immunity is to be achieved.^3^ The public’s perception of who should be prioritised beyond the current list remains unknown but could inform health policy that builds public trust by considering these preferences.

## Methods

This study was approved by the Institutional Review Board of Imperial College Healthcare National Health Service (NHS) Trust (ICHNT). Participants were informed their data would be anonymised for analysis and were free to opt out. We used the NHS’s largest personal health record, the nationwide ICHNT Care Information Exchange (CIE) ^4^, to determine attitudes to the government’s prioritisation list, following the announcement of the second effective vaccine. CIE registrants were sent an email notification to complete a questionnaire within their record (13th November 2020).

Question items determined intent to receive a Covid-19 vaccine in order to refine established modelling^3^ of the target metrics for achieving herd immunity. Further question items focused on attitudes on the UK government’s prioritisation list; and which groups should be prioritised after the initial list has been completed. Descriptive statistics are reported alongside adjusted and unadjusted odds ratios (OR). Free-text analysis and quantification of next-priority groups was performed using natural language processing packages (SpaCy and RegEx) in Python (version 3.7).

## Results

Among 9122 respondents (49.4% response rate), 6521 (71.5%) want Covid-19 vaccination, and 880 (9.6%) would refuse. Although 2068 (22.7%) disagree with the government’s order of priority, 6416 (70.3%) were against being able to pay for vaccination. In response to the question of which groups should be prioritised, unrestricted free-text responses (7,838) indicated teachers (988, 12.6%), BAME (837, 10.7%), general key workers (807, 10.3%) children (582, 7.4%), and university students (529, 6.7%). 32.6% were concerned that the priority list makes no reference to Black, Asian and Minority Ethnic (BAME) groups (table).

**Table.** Care Information Exchange questionnaire response, November 15th, 2020.

|  |  |
| --- | --- |
| **Total N (%)** | 9122 (100) |
| **Age Mean (SD)** | 59.7 (13.6) |
| **Age category N (%)** |  |
| 18-29 | 173 (1.9) |
| 30-39 | 749 (8.2) |
| 40-49 | 1123 (12.3) |
| 50-59 | 2065 (22.6) |
| 60-69 | 2606 (28.6) |
| 70-79 | 2017 (22.1) |
| 80+ | 389 (4.3) |
| **Gender** |  |
| Male | 4176 (45.8) |
| Female | 4945 (54.2) |
| Indeterminate | <5 (0.0) |
| **Ethnicity** |  |
| White background | 6210 (68.1) |
| Non-White background | 1256 (13.8) |
| Prefer not to say | 86 (0.9) |
| Missing** | 1570 (17.2) |
| **If a COVID-19 vaccine was available, would you want to have it?** |  |
| Yes | 6521 (71.5) |
| No - because I believe I have health conditions that make it unsafe | 257 (2.8) |
| No - because I don't believe in vaccination | 71 (0.8) |
| No - because I think I have had COVID-19 infection (but not tested) | 88 (1.0) |
| No - because testing has confirmed I have had COVID-19 infection | 68 (0.7) |
| No - other | 396 (4.3) |
| Not sure | 1678 (18.4) |
| Missing | 43 (0.5%) |
| **Do you agree that this order of priorities is correct?\*** |  |
| **Yes** | 6798 (74.5) |
| **No** | 2068 (22.7) |
| **Missing** | 256 (2.8) |
| **Do you think you should be allowed to pay to have a COVID-19 vaccine without delay?** |  |
| **Yes** | 2435 (26.7) |
| **No** | 6416 (70.3) |
| **Missing** | 271 (3.0) |
| **Number 11 on the government's list is "rest of the population (priority to be determined)". Which groups you think should be prioritised next? Response number (%)\*\*\*** | 7838 (100) |
| **Teachers** | 988 (12.6) |
| **Black, Asian and Minority Ethnic (BAME) Groups** | 837 (10.7) |
| **Key workers (general)** | 807 (10.3) |
| **Children** | 582 (7.4) |
| **University Students** | 529 (6.7) |
| **Public Transport Workers** | 441 (5.6) |
| **Hospitality & Retail** | 278 (3.5) |
| **Other** | 3376 (43.1) |
| **Are you concerned that the government list makes no direct reference to Black and Asian and Minority Ethnic groups?** |  |
| **Yes** | 2971 (32.6) |
| **No** | 5823 (63.8) |
| **Missing** | 328 (3.6) |

\*This question included the UK government's provisional prioritisation list for Covid-19 vaccination for participants to review.
\*\*Ethnicity reported by participants in a previous questionnaire, not completed by 17.2% of participants of the 13th November 2020 questionnaire.
\*\*\* Responses analysed in Python (version 3.7). Tokenization using SpaCy's tokenizer followed by rules-based labelling using RegEx and SpaCy's token matcher, with each individual label verified by the authors. Inclusion limited to labels (groups) with >200 responses.

Excluding those uncertain (N = 1678), yearly increase in age (unadjusted-OR: 1.050 (95%CI:1.044-1.055), adjusted-OR: 1.045 (95%CI:1.039-1.050), and female gender (unadjusted-OR 0.415 (95%CI:0.356-0.482), adjusted-OR 0.540 (95%CI:0.461-0.632) strongly increased and decreased vaccine acceptance respectively, such that a yearly increase in age was associated with a 5% increase in likelihood of vaccination. Overall, 64.2% of females would want vaccination and 12.4% would not, compared to 80.8% and 6.5% of males, respectively.

## Discussion

Although the duration of individual protection from the UK’s now active Covid-19 vaccination programme remains unknown, assuming this to be one year, previous modelling by Anderson et al.^3^ indicates that our headline finding of 71.5% uptake is inadequate for progressing towards herd immunity. A vaccine with 90% efficacy would require 90% coverage in the first year, increasing to 100% of the population for 80% efficacy or less. Potentially inadequate therefore, we identify factors to inform the public health messaging to improve uptake: younger people and females, in combination most exposed to Covid-19^5^, should be proactively targeted. That 33% of respondents were concerned that the priority list makes no reference to BAME groups presents a particular challenge to the UK government, which knows this to be a group not only at increased risk but also with established poor uptake of existing vaccination programmes.^6^ Our results indicate not only the priority considerations for public health messaging necessary to increase the inadequate level of intended uptake, but also the public’s concern at omission of reference to BAME groups and a preference for extending the prioritisation list to teachers and BAME groups.

## Data Availability

Imperial College Healthcare NHS Trust is the data controller. The datasets analysed in this study are not publicly available but can be shared for scientific collaboration subject to meeting requirements of the institution's data protection policy.

## Author Contribution

Patrik Bachtiger: study design, data collection, literature review, data analysis, figures, writing

Alexander Adamson: study design, literature review, figures, data analysis, writing

William A Maclean: figures, data analysis, writing

Jennifer K Quint: study design, literature review, data analysis, figures, writing

Nicholas S Peters: study design, data collection, literature review, data analysis, figures, writing

## Notes

### Competing Interest Statement

The authors have declared no competing interest.

### Funding Statement

This work was funded by: Imperial Health Charity, Imperial Biomedical Research Centre of the National Institute of Health Research, British Heart Foundation, Pfizer Independent Grants, NHSX and Rosetrees Foundation. This work is supported by BREATHE - The Health Data Research Hub for Respiratory Health [MC_PC_19004]. BREATHE is funded through the UK Research and Innovation Industrial Strategy Challenge Fund and delivered through Health Data Research UK.

### Author Declarations

The institutional review board (Imperial College Healthcare NHS Trust Data Protection Office) approved this study and advised ethical approval for data analysis and publication was not required. Participants were informed prior to completing responses that these would be anonymised to inform local and national health policy and were free to opt out.

